# Development of the Kunonga framework for operationalising approaches to health inequality and inequity evidence syntheses

**DOI:** 10.1101/2024.11.13.24317273

**Authors:** Tafadzwa Patience Kunonga, Eugenie Evelynne Johnson, Pauline Addis, Elizabeth Westhead, Peter Bower, Barbara Hanratty, Dawn Craig

## Abstract

**Background:** Health inequalities and inequities are shaped by intersecting social determinants and cumulative life-course experiences. However, conventional evidence synthesis methods often lack the conceptual and analytical tools to capture this complexity. This limits their ability to inform inequality and/or inequity-sensitive policy and practice. In response, we developed a methodological framework to support the systematic integration of intersectionality and life-course perspectives into evidence synthesis.

**Methods:** Framework development followed a three-phase process. First, a systematic review identified conceptual and operational limitations in existing synthesis methods that seek to address health inequality and/or inequity. Second, semi-structured discussions were conducted with eight experts in health inequalities and evidence synthesis to elicit practice-relevant insights aimed at addressing these gaps. Third, findings from both phases were synthesised using a modified framework analysis to construct a structured thematic model aligned with key stages of evidence syntheses (protocol development, data extraction and analysis), informing the design of the methodological framework.

**Results:** The Kunonga Framework offers practical tools and methodological guidance for integrating inequality and/or inequity considerations throughout the evidence synthesis lifecycle. It is underpinned by three core principles: (1) distinguishing between health inequality and health inequity to improve conceptual clarity; (2) applying intersectionality to examine how overlapping social disadvantages shape health outcomes; and (3) adopting a life-course perspective to assess how inequalities and inequities emerge, accumulate and evolve over time. The framework provides practical tools, including logic models, intersectionality matrices and life-stage mapping, to support implementation. A case study on ethnic inequalities in palliative care prescribing illustrated the framework’s feasibility and highlighted its potential to enhance analytical depth and policy relevance.

**Conclusions:** The Kunonga Framework advances evidence synthesis methodology by operationalising established theoretical constructs into actionable guidance. It supports reviewers in considering social complexity and changes across the life-course, helping to explain for whom, how, and in what contexts interventions may work. While developed to support inequality and/or inequity-focused reviews, it can also improve the relevance and depth of broader evidence syntheses. Future research should explore its use across different types of reviews and assess how it can be applied using both qualitative and quantitative methods.

## Background

Evidence synthesis involves systematically gathering, evaluating, and integrating existing research findings to provide a comprehensive and robust summary of available evidence on a particular topic [1]. It encompasses a range of methodologies including systematic reviews, meta-analyses and qualitative syntheses, depending on the nature of the research question [1]. Evidence synthesis plays a crucial role in driving evidence-based decision-making processes, particularly as policymakers increasingly direct their attention towards addressing issues of inequality and inequity [2].

Currently, there is no universally accepted definition of health inequality and health inequity, and the terms are frequently used interchangeably [3]. For the purposes of this article, health inequality refers to differences in health status or determinants among individuals or populations, while health inequity specifically denotes unnecessary, avoidable and unjust differences in health [4]. Health inequalities remain a pressing issue globally, with outcome differences persisting across various demographic groups [5].

While evidence synthesis methodologies play a crucial role in informing healthcare policies and interventions, there are significant gaps in how they address the impacts of health inequalities and inequities [6]. As identified by a recently published systematic review, a range of frameworks have been developed to address aspects of equality and/or equity across health and care research [3]. These include, but are not limited to: the World Health Organization (WHO) conceptual framework on the Social Determinants of Health (SDOH), which distinguishes between structural and intermediary determinants [7]; the Health Equity Impact Assessment (HEIA), designed to assess potential differential impacts of policies or interventions [8]; the PROGRESS-Plus framework,^1^ which identifies key social factors contributing to health inequalities and/or inequities [9, 10]; and, more recently, the EQUALSS Guide Multiple framework,^2^ which supports equity priority-setting across domains [11]. However, while these frameworks are useful for identifying relevant socio-demographic factors, they were not originally developed to guide how inequality or inequity should be understood, interpreted or systematically analysed within evidence synthesis processes. This methodological gap was identified in our earlier systematic review,[3] which highlighted the absence of integrated, operational guidance to consistently embed theoretically grounded equality and/or equity principles, such as intersectionality and life-course approaches, across all stages of the evidence synthesis process [3].

Specifically, the review found that factors contributing to inequality and/or inequity were often considered in isolation, with limited attention to how multiple factors intersect to shape health outcomes in complex, compounding ways [3]. Similarly, life-course approaches, which recognise how cumulative exposures and structural disadvantage across an individual’s lifespan influence health trajectories [12], remain underutilised in evidence synthesis. Yet, applying concepts such as intersectionality and life-course perspectives is essential to understanding how inequalities and/or inequities emerge and persist, a challenge often overlooked in current evidence syntheses. Their limited and fragmented application within evidence synthesis constrains the field’s capacity to support inequality and/or inequity-sensitive decision-making.

Building on these gaps, the Kunonga Framework draws on insights from experts in the field to develop a structured, inequality and/or inequity-centred framework offering methodological guidance for evidence syntheses where currently none exists.

### Research Aim

To develop and present a structured approach for embedding theoretically grounded perspectives on health inequality and/or inequity within evidence synthesis.

## Methods

The development of the proposed framework followed a modified three-stage approach, drawing on recommended phases for methodological framework development [13].

### Phase 1: Evidence Mapping and Problem Framing

The first phase, as outlined in the introduction, involved conducting a comprehensive systematic review to assess existing approaches in inequalities/inequities-focused evidence synthesis. The methods and findings of this review have been reported previously [3], establishing a foundation for developing a more inclusive framework tailored to the complexities of health inequalities/inequities within evidence synthesis. Therefore, this paper will not elaborate further on these review findings but instead concentrates on the subsequent phases that build upon these insights to develop the proposed framework.

### Phase 2: Expert knowledge elicitation

Phase 2 involved small-group key informant discussions with experts in health inequalities, inequities, and/or evidence synthesis to generate insights for methods guidance, addressing the gaps identified in Phase 1. While key informant approaches are typically applied in one-to-one formats [14, 15], we adapted the method to small-group settings to encourage interactive dialogue and peer-level reflection. This approach prioritised conceptual depth and targeted the insights of information-rich participants to examine methodological limitations and identify feasible strategies for integrating inequality and/or inequity considerations into evidence synthesis. As this phase involved expert consultation rather than primary research with patients or the public, it was approved by the Newcastle University Ethics Committee without detailed review (Ref: 61394/2023). This phase was reported in line with the 32-item Consolidated Criteria for Reporting Qualitative Research (COREQ) checklist (Supplementary File 1) [16].

#### Selection and recruitment of experts

Experts were selected based on their demonstrated contributions to their respective fields, evidenced by substantial peer-reviewed publications and other scholarly outputs. This criterion ensured that participants brought a depth of expertise and a history of impactful research, thereby enhancing the rigour and credibility of insights gathered for this study [14, 17]. The recruitment process aimed to ensure diversity of perspectives, including individuals from different sectors and countries. Thirteen experts were contacted via email, introducing the project, and inviting them to participate in the workshop. The email outlined the objectives, expected time commitment and the potential benefits of involvement. Interested experts were asked to confirm their participation, and a suitable date for the discussions was selected based on their availability.

#### Pilot workshops

Two virtual pilot sessions were held with members of the Evidence Synthesis Group at Newcastle University (UK) to gather preliminary feedback and advice for the upcoming key informant discussions. These pilot sessions tested the interview content, activities, logistics and overall participant experience to ensure the effectiveness of the planned approach. Feedback from these sessions directly informed the development of the discussion questions used in the key informant discussions:

- What are the challenges or limitations faced in defining and consistently reporting health inequality and/or inequity within current evidence synthesis methodologies?
- What alternative methodologies or approaches could provide a more multidimensional synthesis of evidence on health inequalities/inequity, capturing a broader range of influencing factors beyond single variables like socioeconomic status?

#### Key informant discussions

Three structured small-group discussions were conducted between March and July 2023. The discussions were led by TPK, an experienced evidence synthesis researcher and facilitated by another researcher (PA). Each interview was held online for approximately 90 minutes and involved at least two experts, allowing for dynamic discussions. The sessions began with a five-minute introduction, followed by a 10-minute presentation of key findings from a case study to establish real-world context [3]. Participants then engaged in a 30-minute brainstorming exercise to foster collaborative problem-solving. Another 10-minute presentation of a second case study enhanced learning, followed by another 30-minute brainstorming session [18]. The sessions closed with a summary and conclusion, reinforcing key insights. An outline of the session agenda is presented in Supplementary File 2.

#### Data collection

Sessions were conducted via Zoom and audio-recorded to capture the conversations and discussions. Recordings were accessible only to the two researchers who facilitated the sessions (TPK and PA), who transcribed and anonymised the data for analysis. For confidentiality, each participant received a unique identifier to attribute their quotations, ensuring that personal data protection protocols were maintained [19].

### Phase 3: Synthesis and framework development

Qualitative data from the key informant discussions were analysed using approach adapted from the principles of framework analysis [20]. Drawing on methodological gaps identified in the review conducted in Phase1 [3], an initial coding framework was developed and applied deductively across transcripts. Two researchers (TPK and EW) independently read the transcripts, coded the data manually and organised them thematically to support comparison across topics. The analysis focused on extracting practice-relevant insights to inform the structure and content of the proposed framework. The final coding structure is presented in Supplementary File 3. To ensure practical relevance, the final themes were aligned with the stages of the evidence synthesis process: protocol development, data extraction and analysis, to support direct application by researchers and decision-makers.

## Results of the key informant discussions

### Characteristics of key informants

Of the 13 originally contacted, eight experts participated in the key informant discussions. The final sample included six experts based in the UK and two based in Africa. Participants brought expertise from a range of domains, including public health, health economics, global health, and evidence methodology. Their roles spanned academia, policy advisory and applied research. Table 1 provides an overview of the participants’ areas of expertise and geographic affiliations.

**Table 1:** Characteristics of experts.

| Expert | Area of Expertise | Current Title | Country/Region | Disciplinary Background |
| --- | --- | --- | --- | --- |
| 1 | Evidence synthesis | Professor of Health & Wellbeing Evidence | UK | Public Health / Evidence Synthesis |
| 2 | Evidence synthesis | Professor in Evidence Synthesis | UK | Information Science / Systematic Reviews |
| 3 | Health inequality | Professor of Health Inequalities | UK | Health Economics / Data Science |
| 4 | Evidence synthesis | Senior Lecturer in Evidence Synthesis | UK | Public Health / Evidence-Based Practice |
| 5 | Health inequality | Public Health Expert | Africa | Health Promotion / Social Determinants of Health |
| 6 | Health inequality | Health Economist | UK | Health Economics |
| 7 | Health equity | Statistical Advisor | UK | Epidemiology / Public Health |
| 8 | Health inequality and synthesis | Emeritus Professor | Africa | Evidence Synthesis |

### Key methodological challenges identified by experts

The discussions generated insights into the practical and conceptual challenges of integrating inequality and/or inequity considerations into evidence synthesis. Experts reflected on the complexities of applying key principles in diverse contexts and offered methodological strategies to address these challenges.

#### Conceptual and terminological ambiguity

A persistent challenge identified by experts was the lack of conceptual clarity and terminological consistency surrounding the terms health inequality and health inequity. While these distinctions are theoretically established, experts noted that in practice, particularly in secondary data analysis, these terms are often conflated or applied inconsistently, with implications for study inclusion, framing, and interpretation. One expert noted the challenge of distinguishing these concepts when working with pre-existing data.

> “When doing a study of real data, the theoretical difference is not always apparent when using secondary data as to whether it relates to inequality or inequity… but generally one should try to use them correctly.” [Expert 7]

In addition, the meanings of inequality and inequity may vary across languages and cultures, leading to different interpretations and understandings. Translating these terms accurately while maintaining their subtle distinctions can be challenging, potentially resulting in miscommunication and ambiguity.

> “…especially when English is not their first language, the translation of inequality and inequity isn’t as clear as it is in English. Internationally, there is a tendency to use inequity, when they mean inequality.” [Expert 4]

> “In French, they are pretty much interchangeable terms… in Finland, they just use equity.” [Expert 3]

Even within the same language, different researchers may use the terms interchangeably or use other terms to define similar concepts.

> “…there is another term that is used quite a lot, particularly in North America, and that is disparity.” [Expert 8]

The implication is that lack of definitional clarity not only undermines analytic precision but risks misclassification of studies, inconsistent application of inclusion criteria and miscommunication within international collaborations. Experts advocated for explicit articulation of these terms at the protocol stage, alongside guidance on how to operationalise them within synthesis.

#### Operationalising intersectionality in evidence synthesis

Experts expressed widespread support for intersectionality as a necessary theoretical lens to understand how structural disadvantage accumulates across multiple dimensions. However, they also emphasised the disconnect between this conceptual ambition and the methodological realities of synthesis. Key barriers included the absence of disaggregated primary data on key socio-demographic factors (e.g. ethnicity, gender identity and socioeconomic status), lack of transparency in subgroup reporting, and limited statistical power for intersectional subgroup analysis.

> “…and if it’s not being collected, it’s not being reported, and you can’t really consider it as a reviewer.” [Expert 2]

Importantly, experts linked these limitations to upstream challenges in primary research design and data governance. For example, one expert highlighted how data protection concerns may preclude collection of variables critical to intersectional analysis.

> “Basically, I don’t think you could gather data from a study what I know. You can’t gather data in the hope that future studies might be able to access that data” [Expert 6]

Another identified tension was the lack of shared understanding about whether intersectionality is a statistical approach (e.g. interaction testing) or a critical interpretive lens. While some experts equated it with methodological tools (e.g. multivariable regression), others emphasised its epistemological role in reshaping what questions are asked, which populations are visible, and how absence of evidence is interpreted.

> “You know, it’s not just one factor which is causing that particular outcome that there are other factors that may need to play, interact and so having that lens in itself is a useful thing” [Expert 1]

One expert noted that qualitative research has an important role to play in addressing these challenges, particularly where quantitative data is insufficient. Qualitative evidence can surface the lived experiences of intersecting disadvantage, give voice to underrepresented groups, and offer unique insights into mechanisms of inequality and/or inequity that are not easily captured through standardised data.

#### Limited integration of life-course perspectives

The third challenge related to the limited integration of a life-course perspective in most evidence synthesis work. Experts observed that many reviews are shaped by the availability of cross-sectional data, which tends to focus on immediate factors affecting health. As a result, it can be difficult to capture how disadvantage builds up over time. This limitation was seen as restricting the ability of syntheses to explore the deeper, long-term pathways that contribute to health inequalities and/or inequities:

> “…because if you’re showing that the resources, for example available to black children in the USA is on average far less…then you see that no, it’s not, it’s not genetic, it’s a lack of resources at the in the very earliest days of life. You look at maternal or child mortality and so on. You can go right back to the start and then follow it through to adult life…” [Expert 8]

The limited use of a life-course perspective was seen as a missed opportunity to understand how health outcomes are shaped by experiences across a person’s life. A more holistic view, tracing key transitions from childhood to adulthood, was seen as essential.

> “… what you need to do is take a holistic approach to social determinants… try to identify factors across the life-course.” [Expert 5]

Experts also linked the life-course perspective to proportionate universalism, suggesting that individuals who have experienced greater disadvantage over time may require tailored or intensified interventions. However, few reviews were seen to incorporate this principle systematically.

> “… and that, I think, brings me to a term…which is I think even more controversial than the term inequity, which is proportionate, universalism which means if someone has had a lifetime of deprivation… you need greater effort to have an effect on their health outcomes.” [Expert 6]

## The Kunonga Framework

In response to the methodological challenges identified, we developed the Kunonga Framework to support the operationalisation of health inequality and inequity considerations in evidence synthesis. Table 2 presents the core components of the Kunonga Framework across key stages of the review process, offering practical guidance to improve conceptual clarity and the integration of intersectionality and life-course perspective.

**Table 2:** The Kunonga Framework: operationalising inequality and/or inequity-focused approaches in evidence synthesis.

| Stage | Component | Practical Guidance |
| --- | --- | --- |
| Protocol Stage | Define Key Terms and Scope | - Explicitly state whether the review focuses on inequality, inequity, or both.<br>- Provide clear definitions and examples to support inclusion/exclusion criteria.<br>- Clarify the conceptual focus to guide data extraction and interpretation.<br>- Involve stakeholders and public contributors early to reflect lived experience. |
|  | Frameworks and Theoretical Approach Selection | - Identify and justify the relevant framework that will guide the review (e.g. PROGRESS-Plus, SDOH, HEIA, EQUALSS GUIDE Multiple) [7-9, 11].<br>- Ensure alignment between framework, definitions, and review objectives.<br>- Justify use of intersectionality and/or life-course approaches depending on the review scope. |
|  | Developing a Logic Model | - Create a visual pathway of how intersecting social determinants and temporal factors influence health outcomes [23].<br>- Identify moderators (e.g. gender, policy context) and mediators (e.g. health literacy, access to care) [24]<br>- Use to guide extraction strategy, subgroup selection, and theory-driven synthesis. |
| Data Extraction Stage | Mapping Data | - Capture disaggregated data on social stratifiers (e.g. ethnicity, gender, or disability) and contextual variables.<br>- Use an intersectionality informed extraction table to record how characteristics are combined or analysed together.<br>- Extract presence/absence of subgroup reporting and interpretation (e.g. structural vs individual explanations).<br>- Map life-course variables: age at exposure, timing of interventions, key transitions (e.g. retirement, caregiving) [29].<br>- Record missing data or absence of stratifiers as analytically meaningful. |
| Analysis Stage | Intersectionality | - Examine overlapping identities and power structures (not just additive factors).<br>- Where possible, apply subgroup meta-analysis or regression models with interaction terms.<br>- Use MAIHDA or IPD where data allow for advanced intersectional analysis [30, 31].<br>- In qualitative or mixed-methods reviews, synthesise narrative findings to examine intersecting disadvantage. |
|  |  | - Note the depth of intersectionality in included studies (descriptive vs analytical use). |
|  | Life-course Analysis | - Code life stages (e.g. early childhood, midlife, old age) to explore cumulative and transitional effects [29].<br>- Apply life-course models [29]:<br>a) Critical period model (early life impact)<br>b) Accumulation model (sustained disadvantage)<br>- Assess timing of exposure and outcome measurement.<br>- Reflect on evidence gaps in temporal coverage (e.g. midlife underrepresented).<br>- Use qualitative insights (e.g. life narratives, contextual framing, timing of exposures) can be used to supplement missing quantitative data. |

### Protocol stage

#### Define key terms and scope

In addition to standard scope-setting practices, this stage requires that reviewers clarify whether the primary focus is on health inequality and/or health inequity, establishing explicit definitions and examples to guide the eligibility criteria. This approach complements existing frameworks by adding specificity, thereby enhancing alignment with the review’s objectives and clarifying priorities for analysis [10]. This stage should include structured involvement of stakeholders, clinical experts and public contributors, particularly those from structurally disadvantaged or underrepresented groups. Early engagement helps ensure the review reflects diverse lived experiences and social contexts [21]. It supports the identification of meaningful inequalities and/or inequities, relevant subgroups, culturally sensitive language, and an ethically grounded analytical lens. This aligns with inclusive research guidance such as the National Institute for Health and Care Research – Innovations in Clinical Trial Design and Delivery for the Under-served (NIHR-INCLUDE) [22].

#### Frameworks and theoretical approach selection

At this stage, reviewers should identify and justify the equity framework or theoretical model that will guide their review. Depending on the review’s scope, focus and population, this may include established tools such as the SDOH, PROGRESS-Plus, HEIA, or the EQUALSS GUIDE Multiple framework for setting equality and/or equity priorities [7–9, 11]. The chosen framework should align with whether the review addresses health inequality, health inequity, or both, and inform eligibility criteria, data extraction and analysis plans.

#### Developing a logic model

Developing a logic model at the protocol stage offers a structured and transparent way to integrate equality and/or equity considerations throughout the synthesis process [23]. Rather than functioning as a static illustration, the logic model should be treated as a dynamic analytical tool that links theory to empirical strategy. It maps the hypothesised pathways through which social identities (e.g. gender, or ethnicity), structural determinants (e.g. housing insecurity, policy environment), and temporal exposures across the life-course (e.g. early-life adversity, transition into retirement) shape differential health outcomes and responses to interventions [23]. These factors may play different analytical roles within the model. Specifically, some may act as moderators, influencing the strength or direction of the relationship between an intervention and an outcome (e.g. socioeconomic status altering the effectiveness of a prevention programme). Others may function as mediators, explaining the mechanisms through which an intervention leads to an outcome (e.g. trust or health literacy mediating the link between provider communication and adherence) [24]. The same factor may play different roles depending on the review context. Articulating these hypothesised relationships early, the logic model may help reviewers make informed decisions about inclusion criteria, subgroup analyses, and what data should be extracted.

## Data extraction stage

The data extraction process must move beyond descriptive cataloguing of socio-demographic characteristics and towards actively capturing how these characteristics intersect, how mechanisms of inequality and/or inequity operate, and how broader contextual factors influence observed outcomes. We recommend that reviewers develop an intersectionality-informed extraction table, in addition to standard data extraction procedures, to systematically capture factors related to inequality and/or inequity. This approach is grounded in the foundational work on intersectionality and its application in health equity research [25, 26]. The table could include a range of core domains. First, reviewers should extract whether social stratifiers (e.g. from SDOH, PROGRESS-Plus, HEIA, or the EQUALSS GUIDE Multiple) are reported in primary studies and, crucially, whether these are analysed in isolation or in combination (e.g. low-income older migrants) [7–9, 11]. Second, reviewers should extract whether outcomes are reported in a disaggregated manner for relevant subgroups or intersectional combinations and whether statistical interaction terms are used to explore compounded or synergistic effects. The interpretive treatment of observed differences could also be recorded, that is, whether the study authors contextualise differences in terms of systemic or structural inequities (e.g. access, discrimination, policy context), or attribute them to individual behaviours or cultural norms [27]. Where logic models have been developed at the protocol stage, they should inform the extraction of variables hypothesised to function as moderators or mediators [24]. Contextual information, including the type of health system, geographic region, and socio-political environment, should also be extracted, as these features are likely to shape both intervention effects and the experience of inequality [28]. A life-course perspective could also be integrated at this stage, through the extraction of timing variables (e.g. age at exposure, intervention duration) and transitional life events (e.g. retirement, caregiving responsibilities) [29]. Finally, reviewers should record patterns of underreporting or omission (e.g. absence of religion), as these gaps are analytically important and should inform later stages of synthesis. Where disaggregated data or statistical estimates are unavailable, reviewers should extract narrative descriptions or author observations to ensure such studies can still contribute to the synthesis. Using interpretation and context-based reasoning in these cases can still offer valuable insights into how and why outcomes vary, instead of leaving those studies out [28]. This approach aligns with calls to systematically extract contextual and mechanistic detail, enabling reviewers to ask not just what works, but for whom, in what context, and why [28]. A template of this table is provided in Supplementary File 4. Reviewers are encouraged to adapt the template to fit the nature of their review. Used effectively, this approach should allow reviewers to synthesise findings not only across different identities, but also across the structural conditions that produce and sustain inequity.

## Analysis stage

### Intersectionality

To support intersectionality-informed analysis, reviewers should avoid examining social factors in isolation or treating them as simply additive. Instead, they should explore how these factors interact and overlap to shape health outcomes in more complex and meaningful ways [25]. Where quantitative data allow, reviewers may conduct subgroup meta-analyses comparing outcomes across different intersections or apply meta-regression to examine whether study-level characteristics such as participant demographics moderate intervention effects. More advanced methods such as Multilevel Analysis of Individual Heterogeneity and Discriminatory Accuracy (MAIHDA) offer a means of modelling between and within group variation across intersecting identities while maintaining statistical robustness [30]. This is particularly valuable because it preserves within-group variability and avoids the pitfalls of dichotomous subgrouping, allowing more accurate estimation of compounded disadvantage [30]. Where individual participant data (IPD) are available, reviewers can model interaction terms directly to test whether outcomes vary significantly across intersectional groupings (e.g. comparing low-income ethnic minority women with high-income white men) [31]. These methods can help reveal whether the combined effect of multiple social factors differs from their individual effects [25].

In cases where such data are not available, reviewers should adopt a qualitative or interpretive synthesis approach. This may involve thematic analysis-based comparisons across identity intersections, drawing on both narrative findings and contextual data. Particular attention should be paid to how primary studies explain observed differences, specifically whether they rely on behavioural, cultural or biological explanations, or whether they acknowledge structural determinants such as discrimination, racism, class inequality or exclusion from services [32]. Reviewers should also reflect on the depth of intersectionality in each study, considering whether it is used merely as a demographic descriptor or whether it meaningfully informs the study’s theoretical framework and analytical approach.

### Life-course analysis

Reviewers should consider incorporating a life-course perspective to examine how exposures, interventions and outcomes are distributed, sequenced and accumulated across different life stages. Syntheses can be structured to identify whether and how evidence addresses critical periods (e.g. early childhood), cumulative disadvantage (e.g. long-term poverty), or transitional stages (e.g. entry to the labour market, retirement). This can be operationalised by coding for the life stage targeted by each study and mapping outcomes accordingly [29]. Where data permit, reviewers may apply age-stratified meta-analyses, trajectory modelling or synthetic cohort approaches to examine when inequalities emerge, persist or widen over time [29]. Life-course models such as the critical period framework, which emphasises the lasting impact of early-life exposures, and the accumulation model, which highlights the additive effects of repeated or prolonged disadvantage across time, can guide interpretation by highlighting the long-term impact of early exposures and the additive effects of disadvantage, respectively [12]. Reviewers should also reflect on evidence gaps in terms of temporal coverage. If the evidence base is disproportionately concentrated in early childhood and late life, with minimal attention to mid-life or transitional periods, this may limit the applicability of findings to cumulative or long-term policy responses.

## Application of the Kunonga framework: a case example

To demonstrate the application of the Kunonga Framework, we present a case study on its integration into the conduct of a rapid systematic review on ethnic inequalities in palliative care prescribing in high-income countries [33]. Although the review protocol had been registered prior to the formal publication of the framework, its conceptual components were used to inform the review’s design, data extraction, analysis and interpretation. This allowed us to assess the feasibility and added value of applying the framework in practice. Table 3 summarises how the Kunonga Framework was operationalised, highlighting key insights and methodological challenges encountered during the review process.

**Table 3:** Application of the Kunonga framework in a review of ethnic inequalities in palliative prescribing.

| Framework Stage | Component | Application in Case Study | Insights and Challenges |
| --- | --- | --- | --- |
| Protocol | Define Key Terms and Scope | The review focused on health inequalities, defined as observable differences in palliative prescribing among ethnic groups. | The framework clarified the conceptual distinction between inequality and inequity. Retrospective reflection revealed how explicit definition at protocol stage improves alignment between aims, eligibility criteria, and analytical depth. |
|  | Frameworks and Theoretical Approach | PROGRESS-Plus was adopted post protocol registration to guide identification of relevant social stratifiers in data extraction and analysis [9]. | The Kunonga Framework encouraged inclusion of life-course and intersectional perspectives, but original protocol did not anticipate these. |
|  | Logic Model | A logic model was developed post protocol to clarify hypothesised pathways linking social determinants (e.g. ethnicity, age) to prescribing outcomes. Moderators from PROGRESS-Plus were included [9]. | Despite being developed post hoc, the model helped make explicit the mechanisms and assumptions underlying observed inequalities |
| **Data Extraction** | Mapping Data | Data extraction was reorganised to reflect intersecting demographic and social variables (e.g. age, gender, SES, place of residence, ethnicity), and life-course elements such as age at diagnosis. | The framework supported a more unique categorisation of data, but limitations in primary study reporting constrained the extent of meaningful intersectional or temporal analysis. |
| **Analysis** | Intersectionality | Analysis considered multiple PROGRESS-Plus factors (e.g. ethnicity, income, age) [9], using both adjusted regression models and subgroup comparisons where available. | The framework revealed analytical gaps: few studies tested for interaction effects or explored intersecting inequalities. Narrative synthesis and author commentary were used to supplement quantitative findings. |
|  | Life-course Perspective | Limited application - only one study explored age-specific patterns in prescribing. Most lacked data to assess how prescribing varied over the life course or in relation to illness progression. | The framework exposed significant gaps in temporal reporting. Its application highlighted the need for longitudinal or stage-specific data to enable assessment of cumulative disadvantage or life-stage-specific barriers. |

Key: PROGRESS-Plus: Place of residence, Race/ethnicity, Occupation, Gender, Religion, Education, Socioeconomic status, Social capital, plus additional factors such as personal characteristics, features of relationships, and time-dependent relationships.

### Protocol Stage

#### Define key terms and scope

The review focused on health inequalities rather than inequities because we were specifically interested in identifying and describing measurable differences in palliative care prescribing among ethnic groups. Health inequality was defined as observable differences, such as variations in medication type, dosage, or frequency of pain relief, without assuming these differences were unfair or avoidable [33]. This focus aligned with the review’s aim to document variations in prescribing practices in a way that was empirically grounded and descriptively clear [33]. Assessing inequities would require a different evidentiary and interpretive approach, incorporating ethical and contextual analysis that was beyond the scope of this review.

#### Frameworks and theoretical approach selection

The review adopted the PROGRESS-Plus framework after protocol registration to identify and extract factors relevant to prescribing inequality [9]. It offered a pragmatic and transferable structure for identifying key factors relevant to inequality and supported a systematic approach to data extraction.

#### Developing a logic model

A logic model was developed to clarify hypothesised pathways linking palliative prescribing practices to inequality-relevant outcomes (Fig. 1). The model articulated key assumptions, intervention components and expected short-, medium- and long-term outcomes. Moderating factors were drawn from PROGRESS-Plus factors to highlight how social position and identity-based disadvantage might influence prescribing patterns and symptom management [9].

**Figure 1:**
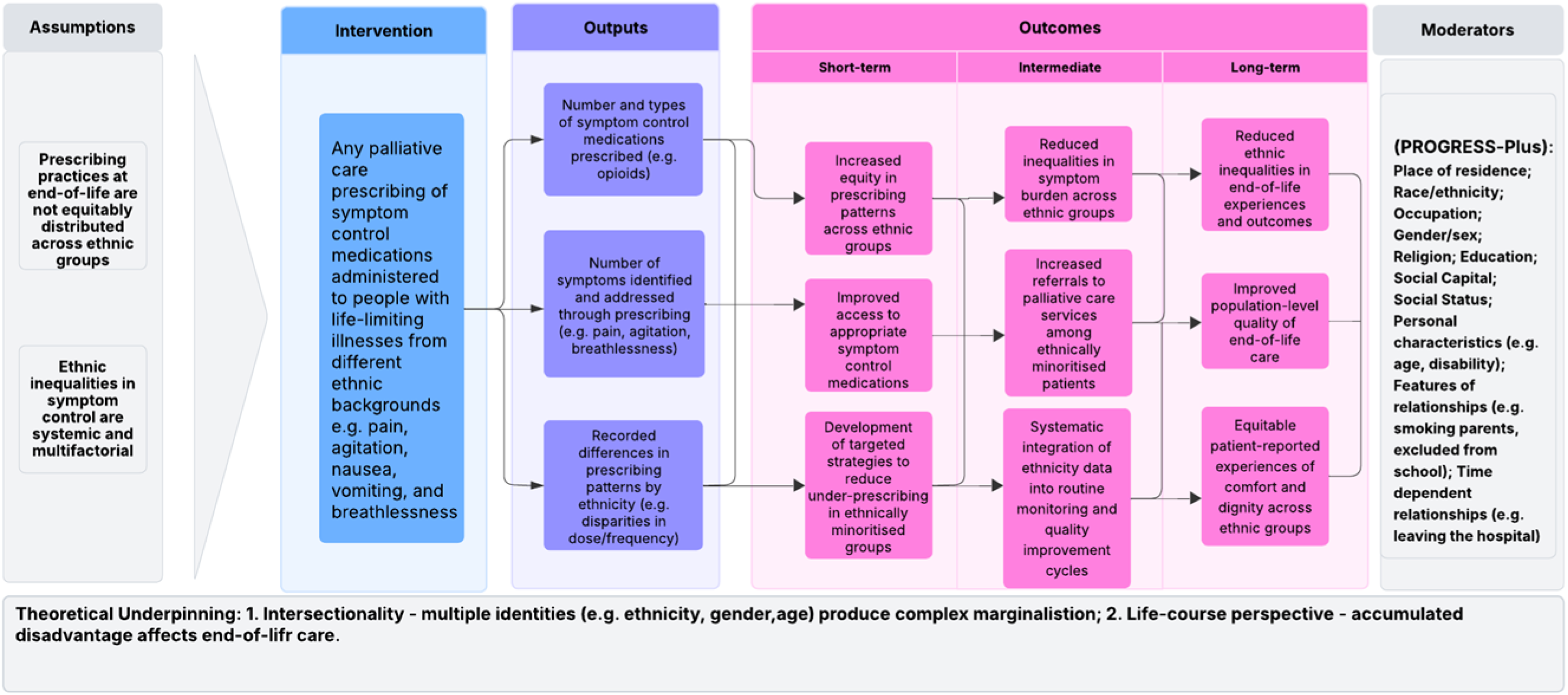
Logic model of ethnic inequalities in palliative prescribing: integrating intersectionality and life-course perspectives.

## Data extraction stage

Data were extracted using a structured Excel form capturing study characteristics, participant demographics, prescribing practices and reported moderating factors. Guided by the Kunonga Framework, the extraction process was designed to highlight intersecting social and demographic factors relevant to health inequalities, including age, gender, socioeconomic status, place of residence and ethnicity. The data were also organised to capture life-course dimensions, such as age at diagnosis and experiences of persistent poverty [33].

## Analysis and synthesis stage

### Intersectionality

Applying an intersectional lens during analysis was challenging due to limited data disaggregation within the included studies. However, we systematically examined how multiple PROGRESS-Plus factors were addressed individually and in relation to ethnicity [9, 33]. In addition to extracting disaggregated findings, we assessed whether studies used statistical approaches that accounted for multiple social determinants simultaneously. This included identifying the use of multivariable regression models that adjusted for intersecting factors (e.g. ethnicity, income, age) in the analysis of access to palliative care or symptom management [33]. While these models did not always test for interaction effects explicitly, they offered partial insights into how multiple axes of inequality were considered concurrently. Where studies used stratified analysis or subgroup comparisons (e.g. by ethnicity or deprivation status), we examined whether findings varied systematically across these groups [33]. However, only a minority of studies explicitly considered the interplay between demographic factors or reported subgroup-specific outcomes in a way that allowed for intersectional interpretation. To address these gaps, we supplemented the synthesis with narrative insights, drawing on contextual information and author commentaries where intersecting disadvantages were acknowledged but not explored [33].

### Life-Course Analysis

Incorporating a life-course perspective into the analysis was similarly constrained by the limitations of the primary studies. Only one study reported age-specific trajectories or explored how prescribing practices varied across the stages of ageing, chronic illness progression or proximity to end of life. This lack of temporal granularity limited our ability to assess whether observed inequalities reflected cumulative disadvantage or stage-specific barriers to care [33].

## DISCUSSION

### Main findings

This study introduces the Kunonga Framework as a structured, operational approach to embedding equality and/or equity-sensitive principles, specifically intersectionality and life-course theory, into evidence synthesis. While the theories are well-established in public health and social science literature [12, 26, 34], their practical application within evidence synthesis has remained inconsistent and underdeveloped. The Framework addresses key methodological gaps that were first identified in a prior systematic review [3], and subsequently explored through expert discussions. These gaps include the persistent conflation of health inequality and inequity, the limited integration of intersectionality due to insufficient data disaggregation and reliance on single-axis analyses, and the underuse of life-course perspectives, often constrained by a lack of temporally sensitive or longitudinal data [4, 29, 35, 36]. The framework offers practical guidance across the synthesis process, from clarifying conceptual definitions and selecting equality and/or equity-focused tools at the protocol stage (e.g. the SDOH, PROGRESS-Plus, HEIA, or the EQUALSS GUIDE Multiple framework), to supporting the identification of demographic and social factors and extracting and analysing data through an intersectional and life-course lens [7–9, 11].

### Strengths and limitations

This study used a structured, three-phase approach to develop the Kunonga Framework, integrating inequality and inequity considerations into evidence synthesis. The framework’s construction followed an iterative process: a systematic review to identify gaps; key informant discussions with eight health inequality and inequity experts; and synthesis of findings into a practical framework. This phased design allowed for continual refinement, ensuring that each phase built upon previous insights to produce a comprehensive, context-sensitive framework.

One key strength of this approach is its methodological rigour. The systematic review phase established a solid foundation by identifying gaps in existing inequality/inequity-focused evidence synthesis methodologies, ensuring that the framework built upon, rather than duplicated, prior work [3]. The use of key informant discussions provided qualitative insights, contextualising these gaps and guiding framework elements by capturing unique perspectives on the needs and challenges of inequality and/or inequity-focused syntheses [14, 37]. This combined approach facilitated a holistic understanding of the multi-dimensional factors influencing health inequalities, which is crucial for developing effective interventions. The use of an adapted framework analysis allowed us to work from a priori concepts while remaining open to emergent themes. By structuring the analysis around stages of the evidence synthesis process (protocol development, data extraction and analysis), the framework offers clear guidance for researchers seeking to embed inequality and/or inequity equity considerations in a consistent manner.

However, there are limitations in the process of constructing the framework. First, the framework’s reliance on a limited number of experts means it may not fully represent the diverse perspectives and contexts relevant to global health inequities [38, 39]. While this offered depth in terms of specific expertise, the exclusion of patients, public contributors and voluntary or charitable organisations constrained the diversity of voices and experiential knowledge captured [21]. Including lived experience perspectives may have revealed different priorities, inequality and/or inequity concerns or framings, particularly around what counts as meaningful inequalities or inequities, or how intersecting disadvantage can manifest. Third, while the framework was tested using a single case study, it has not yet been implemented across diverse review designs or topics.

### Implications for research

The development of the Kunonga Framework marks a step toward operationalising inequality and/or inequity-sensitive methodologies in evidence synthesis. However, its introduction also highlights several critical areas for future research. First, while the framework offers structured guidance for integrating intersectionality and life-course perspectives, its practical application remains underexplored across diverse evidence synthesis types and health contexts. Future studies should prospectively apply the framework to assess its adaptability, usability and impact on review outcomes. Second, the framework’s emphasis on intersectionality and life-course theory exposes persistent limitations in the primary literature, particularly the lack of disaggregated and longitudinal data. These gaps constrain the ability of reviewers to conduct unique analyses and highlight the need for upstream improvements in study design and reporting. Research funders and institutions should prioritise data infrastructure that supports inequality and/or inequity-focused synthesis, including the routine collection of sociodemographic variables and life-course indicators. Third, while the current recommendations are primarily qualitative and interpretive in nature, there is a need to explore quantitative methods for incorporating intersectionality into evidence synthesis. This includes the development of approaches capable of modelling complex, interacting dimensions of structural disadvantage in ways that are theoretically grounded and statistically robust. Fourth, although developed to support reviews focused on health inequality and inequity, the Kunonga Framework can also be used in broader evidence syntheses where understanding differences across groups, contexts, or time is important.

Finally, the application of the Kunonga Framework to a rapid review on ethnic inequalities in palliative care prescribing provides an initial demonstration of its feasibility and added value [33]. The case study illustrates how the framework can enhance conceptual clarity, guide data extraction and support intersectional and temporal analyses, even when applied following protocol registration. Future research should build on this example by embedding the framework prospectively in review protocols, thereby strengthening its empirical foundation and refining its components through iterative testing.

### Implications for policy

The Kunonga Framework will not only enhance our comprehension of health inequalities or inequities within evidence syntheses but will also offer a pathway for translating research findings into actionable policy recommendations and interventions. By identifying the root causes of health inequalities/inequities and understanding their dynamic nature across the life-course and among diverse populations, policymakers and practitioners will be better able to design targeted interventions that address underlying structural factors, but also to make decisions that are informed by a clearer understanding of potential inequalities and/or inequities.

## Conclusion

This paper contends that the current state of evidence synthesis in health inequality and inequity research is inadequate to capture the multifaceted and dynamic nature of the issue. The Kunonga Framework offers a structured, theory-informed approach for integrating health inequality and/or inequity considerations into evidence synthesis. By embedding principles of intersectionality and the life-course perspective, it responds to the growing demand for equity-sensitive methodologies that move beyond surface-level subgroup analysis. The framework provides practical guidance across the review lifecycle.

Although the framework was developed to support inequality and/or inequity-focused reviews, its components are equally useful in broader evidence syntheses where variation across populations, contexts, or stages of life may influence how interventions work. As inequality and inequity remain pressing concerns for health and care policy, the Kunonga Framework can help ensure that reviews are more inclusive, reflective, and relevant to real-world decision-making. Future research should test the framework in prospective reviews, assess its use with both qualitative and quantitative methods, and explore its adaptability across different review designs and topics.

## Supporting information

Supplementary File 1 COREQ_Checklist

Supplementary File 2 Key Informant Discussions Agenda

Supplementary File 3 Analysis Coding Structure

Supplementary File 4 Intersectionality- informed data extraction template for evidence synthesis

## Data Availability

All data relevant to the study are included in the article or uploaded as supplemental information.

## Declarations

### Ethical approval

Ethical approval for the study was granted by the Faculty of Medical Sciences Research Ethics Committee at Newcastle University, United Kingdom (Reference: 61394/2023). All methods were carried out in accordance with relevant institutional guidelines and regulations. As this study involved professionals acting in their expert capacity, and did not include patients, service users, or members of the public, the study was deemed exempt from full ethical review.

### Consent for publication

Informed consent was obtained from all participants prior to participation.

### Availability of data and materials

All data generated or analysed during this study are included in this published article [and its supplementary information files].

### Competing interests

The authors declare that they have no competing interests.

### Funding/support

This research is funded through the National Institute for Health and Care Research (NIHR) Policy Research Unit in Older People and Frailty (funding reference PR-PRU-1217-2150). As of 01.01.24, the unit has been renamed to the NIHR Policy Research Unit in Healthy Ageing (funding reference NIHR206119). The views expressed are those of the author(s) and not necessarily those of the NIHR or the Department of Health and Social Care.

### Author contributions

TPK conceptualised the study, developed the methodology, curated the data, prepared the original draft, acquired resources, and managed project administration. TPK, BH, PB, and DC contributed to visualisation, investigation, and validation. TPK and EW performed formal analysis. BH, PB, and DC supervised the project. TPK, EEJ, PA, EW, BH, PB, and DC reviewed and edited the manuscript.

## Acknowledgements

I would like to thank my PhD internal assessors, Professor Sheena Ramsay and Professor Thomas Scharf from Newcastle University, and members of the expert panel who generously provided knowledge and expertise.

## Patient involvement

No patients or public were involved in the planning or drafting of this article.

## Supplementary materials

Supplementary File 1: Consolidated Criteria for Reporting Qualitative Research (COREQ) checklist.

Supplementary File 2: Key Informant Discussions Agenda.

Supplementary File 3: Analysis Coding Structure.

Supplementary File 4: Intersectionality-informed data extraction template for evidence synthesis.

## List of abbreviations

COREQ: Consolidated Criteria for Reporting Qualitative Research
EFW: Equity-Focused Workshops
EQUALSS GUIDE Multiple framework: Ethnicity and race, Qualifications and education, Underserved area, Age, Language and religion, Sex, Sexual orientation, Gender identification, Underrepresented groups (inclusion groups), Income and wealth, Disability (physical, mental and learning), Employment and occupation, and Multiple disadvantages.
HEIA: Health Equity Impact Assessment.
MAIHDA: Multilevel Analysis of Individual Heterogeneity and Discriminatory Accuracy.
NIHR-INCLUDE: National Institute for Health and Care Research Innovations in Clinical Trial Design and Delivery for the Under-served
PRISMA Equity: Preferred Reporting Items for Systematic Reviews and Meta-Analyses-Equity.
PROGRESS Plus: Place of residence, Race/ethnicity, Occupation, Gender, Religion, Education, Socioeconomic status, Social capital, plus additional factors such as personal characteristics, features of relationships, and time-dependent relationships.
SDOH: Social Determinants of Health.

## Footnotes

1 PROGRESS-Plus: Place of residence, Race/ethnicity/culture/language, Occupation, Gender/sex, Religion, Education, Socioeconomic status, and Social capital, plus additional factors such as personal characteristics (e.g., age, disability), relational influences (e.g. smoking parents), and time-dependent variables (e.g. hospitalisation).

2 EQUALSS GUIDE: Ethnicity and race, Qualifications and education, Underserved area, Age, Language and religion, Sex, Sexual orientation, Gender identification, Underrepresented groups (inclusion groups), Income and wealth, Disability (physical, mental and learning), Employment and occupation, and Multiple disadvantages.

## Notes

### Competing Interest Statement

The authors have declared no competing interest.

### Summary of Updates

Title revised to reflect operational focus of the framework; references to additional frameworks added, including WHO SDOH, HEIA, and EQUALSS GUIDE Multiple; methods section restructured into three phases (evidence mapping, expert elicitation, and synthesis); description of expert input updated to reflect small-group discussions; key informant interviews reframed as expert discussions; framework analysis cited to correctly reflect the analytical approach used in the study; case study on ethnic inequalities in palliative care prescribing added to demonstrate framework application and feasibility; author list updated to include Elizabeth Westhead; author contributions and affiliations revised.

