## Supplementary File 2 Key Informant Discussions Agenda for "Development of the Kunonga framework for operationalising approaches to health inequality and inequity evidence syntheses"

| **Date** | xxxxx | |
| --- | --- | --- |
| **Time** | xxxxx | |
| **Location** | Via MS Teams Video Conferencing: | |
| **Chair** | PK | |
| **Facilitator** | PA | |
|  | **Agenda** | **Who** |
| 1. | Welcome and housekeeping (10mins) | **PA** |
| 2. | Aims of the meeting (5mins) | **PK** |
| 3. | Presentation of key findings from [A systematic review finds a lack of consensus in methodological approaches in health inequality/inequity focused reviews](https://www.sciencedirect.com/science/article/pii/S0895435623000343) (10mins) | **PK** |
| 4. | Brainstorm exercise (20 – 30mins)  **Key finding:** The analysis found that there was inconsistency in the way inequality or inequity is reported in evidence synthesis. Various terms were used to describe the focus of the reviews (e.g. equity, inequality, inequity), but they were seldom defined and sometimes used interchangeably.  **Question:** What are the challenges or limitations faced in defining and consistently reporting health inequality and/or inequity within current evidence synthesis methodologies? | **All** |
| 5. | Break (5 mins) |  |
| 6. | Presentation of case study based on key findings from [Health interventions and the unseen impact on equality](https://doi.org/10.1016/S2666-7568(22)00268-9) (10mins) | **PK** |
| 7. | Brainstorm exercise (20 – 30mins)  **Key finding:** Health inequality/inequity research tends to be one-dimensional e.g. focus on SES as in the case study. However, it is possible that other factors influence disability free life expectancy such as gender and individual health behaviours.  **Question:** What alternative methodologies or approaches could provide a more multidimensional synthesis of evidence on health inequalities/inequity, capturing a broader range of influencing factors beyond single variables like socioeconomic status? | **All** |
| 8. | Summary of discussions (5mins) | **PK** |
| 9. | Next steps (5mins) |  |
| 10. | Concluding remarks (5mins) | **PK** |

| **Invited Attendees** |
| --- |
