## Supplementary File 3 Analysis Coding Structure for "Development of the Kunonga framework for operationalising approaches to health inequality and inequity evidence syntheses"

| **Main theme** | **Subtheme** | **Illustrative Quote (with attribution)** | **Interpretive note** |
| --- | --- | --- | --- |
| **Conceptual and terminological ambiguity** | Language and Regional Variability | “…especially when English is not their first language, the translation of inequality and inequity isn’t as clear as it is in English. Internationally, there is a tendency to use inequity, when they mean inequality.” – Expert 4  “…there is another term that is used quite a lot, particularly in North America, and that is disparity.” - Expert 8  "In French, they are pretty much interchangeable terms… in Finland, they just use equity." – Expert 3 | Linguistic and cultural differences affect conceptual clarity and make standardised synthesis more difficult. The variability complicates comparative work across set tings. |
|  | Ideological Barriers | "One barrier could be ideological… a reluctance to accept some differences as unjust or avoidable." – Expert 6 | Political and moral values influence whether differences are interpreted as structural injustices or natural variation. This affects uptake of equity frameworks in practice. |
|  | Conceptual Overlap and Measurement Challenges | "I imagine there are times when it's impossible to sort out if a difference is inequality or inequity… quite a messy area." – Expert 1  When doing a study of real data, the theoretical difference is not always apparent when using secondary data as to whether it relates to inequality or inequity… but generally one should try to use them correctly.” - Expert 7 | Ambiguity arises when differences may appear biologically based but also relate to structural disadvantage, complicating decisions in evidence inclusion and categorization. |
|  | Discipline- and Context-Specific Use | Terms are often used interchangeably… and even within papers, equity and inequality are not always clearly defined." – Expert 2 | The use of terms is inconsistent across academic disciplines and influenced by societal norms, making systematic comparison challenging in synthesis. |
|  | Implications for Review Inclusion | "If you're doing a review and only include papers that say 'inequality', you may wrongly exclude those labelled 'inequity'." – Expert 5 | A strict definition may lead to inappropriate exclusion; interpretive flexibility and careful conceptual scoping are needed in review protocols. |
| **Operationalising intersectionality in evidence synthesis** | Gaps in Primary Data Collection | "If it's not being collected, it's not being reported, and you can't really consider it as a reviewer." – Expert 2 | Lack of primary data collection on factors influencing health outcomes impairs reviewers’ ability to conduct intersectional analyses. |
|  | Ethical and Policy Constraints | “Basically, I don't think you could gather data from a study what I know. You can't gather data in the hope that future studies might be able to access that data” – Expert 6 | Current data governance rules restrict proactive data collection for secondary use, limiting intersectional potential. |
|  | Analytical Lens and Theory | “You know, it's not just one factor which is causing that particular outcome that there are other factors that may need to play, interact and so having that lens in itself is a useful thing." – Expert 1 | Even when full analysis is not possible, applying an intersectionality lens improves interpretive depth and critical reflection. |
|  | Inconsistency in defining Indicators | Some factors have heterogenous definitions e.g. SES – Expert 5 | Ambiguities in indicators reduce comparability across studies and hinder structured intersectional coding. |
|  | Public and Stakeholder Involvement | We could recruit individuals with lived experience, to get their views on some topics – Expert 5 | Where data are missing, involving affected communities may offer experiential insights to inform intersectional considerations. |
| **Limited integration of life-course perspectives** | Cumulative Disadvantage | "Early stresses and subsequent deaths are all cumulative and have an effect later in life." – Expert 8 | Health inequalities/inequities emerge through the accumulation of disadvantage across critical life stages, reinforcing the need for longitudinal framing. |
|  | Origins of Adverse Health | ". . .because if you're showing that the resources, for example available to black children in the USA is on average far less. . .then you see that no, it's not, it's not genetic, it's a lack of resources at the in the very earliest days of life. You look at maternal or child mortality and so on. You can go right back to the start and then follow it through to adult life...” – Expert 8 | Understanding contemporary health inequalities requires tracing their roots across earlier exposures and experiences throughout life. |
|  | Area vs Individual-Level Deprivation | "There is a slight distinction between the poverty of the area they live in and their own poverty." – Expert 5 | Area-level deprivation may compound individual disadvantage and must be distinguished when interpreting long-term impacts on health. |
|  | Holistic Mapping of Social Determinants Across the Life-Course | ". . . what you need to do is take a holistic approach to social determinants. . . try to identify factors across the life-course.” Expert 5 | A fragmented view of exposures misses cumulative, interacting effects. Synthesising evidence with a life-course lens requires connecting diverse social determinants (e.g. housing, education, income) across time, rather than analysing them in isolation or at a single life stage. |
|  | Policy Frameworks and Proportionate Universalism | ". . . and that, I think, brings me to a term. . .which is I think even more controversial than the term inequity, which is proportionate, universalism which means if someone has had a lifetime of deprivation... you need greater effort to have an effect on their health outcomes." – Expert 6 | Proportionate universalism highlights that life-course-informed equity interventions should be scaled according to cumulative disadvantage. |
