## Supplementary File 4 Intersectionality- informed data extraction template for evidence synthesis for "Development of the Kunonga framework for operationalising approaches to health inequality and inequity evidence syntheses"

| **Extraction element** | **Explanation and guidance** |
| --- | --- |
| **Study ID** | Unique identifier (e.g. Author, Year). Enables traceability and cross-referencing. |
| **Reported social stratifiers** | List reported social characteristics (e.g. age, ethnicity, gender, socioeconomic status, disability, religion). Use frameworks like SDOH, PROGRESS-Plus, HEIA, or EQUALSS GUIDE Multiple as reference [7-9, 11]. |
| **Disaggregation level** | Specify stratification granularity:  • None  • Single stratifier (e.g. just age)  • Multiple (non-intersecting) (e.g. age and gender, analysed separately)  • Full intersectional disaggregation (e.g. age × gender × ethnicity).  *Clarify using examples (e.g. “Reported by gender and ethnicity separately”).* |
| **Intersectionality evidence** | Assess analytical depth:  • Descriptive – Lists groups but does not analyse differences  • Analytical – Subgroup or interaction analysis (e.g. regression with interaction terms)  • Interpretive – Narrative integration of intersecting disadvantage (e.g. lived experience, qualitative insights). |
| **Mechanism of inequality/inequity** | What explanation is offered for observed differences?  • Individualised – Attributed to behaviours, attitudes, preferences  • Structural/Systemic – Attributed to racism, policy, discrimination, access barriers.  *Note which type is present, and whether explanation aligns with observed data.* |
| **Moderators / mediators reported** | List any variables identified as:  • Moderators – Modify strength or direction of effect (e.g. gender, education)  • Mediators – Explain the mechanism (e.g. health literacy, trust).  *Refer to logic model if available.* |
| **Life-course markers** | Extract temporal/life-stage dimensions:  • Age at exposure/intervention  • Duration of exposure or disadvantage  • Critical transitions (e.g. retirement, parenthood, illness onset).  *Supports mapping of cumulative or stage-specific effects.* |
| **Temporal framing** | How is time conceptualised? Choose one:  • Cross-sectional  • Longitudinal  • Life-course  • Unspecified  *Useful for judging appropriateness of applying accumulation or critical period models.* |
| **Inequality/inquity interpretation** | Does the study interpret findings through an equity lens?  • No – Differences presented descriptively only  • Implicit – Suggests injustice or need for change without stating explicitly  • Explicit – Labels outcomes as unjust, unfair, or avoidable.  *Support with quotes or author framing where possible.* |
| **Missing or omitted data** | Identify important social dimensions not reported or not analysed, especially where context suggests they should be.  *E.g. “Ethnicity collected but not analysed”; “No data on disability or religion.”*  Supports “absence as evidence” to highlight structural erasures or data limitations. |
| **Contextual factors** | Record broader setting characteristics likely to influence inequality or interpretation:  • Health system type (e.g. universal, insurance-based)  • Policy/legal context (e.g. entitlements, discrimination laws)  • Geographic/cultural setting  *Supports explanatory and realist analysis.* |
| **Data source type** | Note the type of evidence:  • Administrative records  • Surveys  • Clinical trials  • Lived experience/qualitative interviews  *Allows assessment of voice, bias, and data richness for equity-sensitive interpretation*. |
| **Power for subgroup analysis** | Assess whether the study had adequate power for subgroup or interaction analyses. Note if authors reported limitations (e.g. small sample sizes, wide confidence intervals).  *Flags analytic constraints that affect how inequality/inequity can be assessed.* |

Key: SDOH: Social Determinants of Health; PROGRESS Plus: Place of residence, Race/ethnicity, Occupation, Gender, Religion, Education, Socioeconomic status, Social capital, plus additional factors such as personal characteristics, features of relationships, and time-dependent relationships; HEIA: Health Equity Impact Assessment; EQUALSS GUIDE Multiple framework: Ethnicity and race, Qualifications and education, Underserved area, Age, Language and religion, Sex, Sexual orientation, Gender identification, Underrepresented groups (inclusion groups), Income and wealth, Disability (physical, mental and learning), Employment and occupation, and Multiple disadvantages.
